# Development and Validation of a Real-Time PCR for the Detection of *Haycocknema perplexum*

**DOI:** 10.1101/2025.10.03.25336203

**Authors:** Kym Lowry, Bijendra Parmar, Rita Gupta, Thomas Robertson, David Whiley, Patrick NA Harris, Natalie Davidson

## Abstract

*Haycocknema perplexum* is a rare and emerging cause of parasitic myositis. Detection and surveillance are of growing importance given the increasing degree of climate conditions conducive to transmission. In this study, we developed a real-time PCR method for direct detection of two genomic regions of *Haycocknema perplexum*. The real-time PCR assays, SSU-PCR and COX1-PCR targeted the small subunit of nuclear ribosomal RNA and cytochrome oxidase-1 genomic regions, respectively. The performance of the assays was assessed using a panel of *H. perplexum* samples, both fresh frozen and formalin fixed paraffin embedded tissue (FFPE) (n = 22, derived from eight patients) and non-*H. perplexum* (n = 8) tissue biopsy specimens. Both *H. perplexum* assays showed 83% sensitivity and 100% specificity, with negative and positive predictive values of 100% and 93% respectively. The results indicate that the LOD was 10^−5^ dilution (C_t_ value 40.2) for COX1-PCR, and 10^−3^ dilution (C_t_ value 39.6) for SSU-PCR, making the latter a more sensitive assay for detecting lower concentrations of organism in the patient biopsy. The sensitivity of fresh frozen samples was superior to FFPE samples. All but 1 sample was negative following treatment. Feasibility of real-time PCR detection of *H. perplexum* directly from tissue biopsies has been demonstrated for diagnosis, possible test of cure and could enhance transmission surveillance.

## 1. INTRODUCTION

*Haycocknema perplexum* is a rare and emerging cause of parasitic myositis in Australia. It was first described in 1998 in Tasmania from an English botanist with myositis. Since that time, an additional 13 cases have been reported (1). All these cases have been associated with residence or travel to either Northern Queensland or Tasmania in Australia. To date, zoonotic and environmental reservoirs remain unknown; consequently the risk factors for human infection are not clear, resulting in delayed diagnosis and treatment (2).

The organism historically has been considered to be a member of the Robertdollfusidae family of the superfamily Muspiceoidea (3). Other related nematodes have been identified in native Australia marsupials, however, interestingly, a similar organism was identified in a horse in Switzerland (4). Some of the published cases have reported a history of bushmeat ingestion or close contact with native animals with hypothesized routes of transmission including via a direct cutaneous route, via ingestion or via an intermediate arthropod host (5). The life-cycle of the organism can occur within a human cell with efficient auto-reinfection occurring and infecting adjacent myocytes (6).

Clinical findings consistent between the reported cases include progressive weakness often with associated dysphagia and weight loss. Laboratory findings include an elevated creatinine kinase (CK), peripheral eosinophilia and in many cases abnormal liver function tests (1). Diagnosis is challenging and most commonly is confirmed on a muscle biopsy with non-encysted nematodes visualised intracellularly (∼350µm in length and 20µm in width). Treatment includes anti-parasitic therapy with Albendazole and importantly the use of corticosteroid therapy has been associated with worse clinical outcomes (7).

Molecular techniques are not widely available for nematode diagnostics but have been previously put forward as an adjunctive tool for diagnosis as well as to better inform both biology and taxonomy (5, 8). We aimed to collate all available positive samples from Queensland to develop an in-house real-time TaqMan PCR, based on previous work, targeting both the SSU (small subunit of nuclear ribosomal RNA) and COX-1 (cytochrome oxidase-1) genomic regions of *H. perplexum* to determine the performance characteristics of the assay for use on muscle biopsy samples as a confirmatory tool for diagnosis (8). We also aimed to perform gene sequencing of positive samples and subsequent phylogenetic analysis to gain knowledge regarding taxonomy.

## 2. MATERIALS AND METHODS

### 2.1 Samples

The assays were validated using specimens submitted to Pathology Queensland and retrieved from long-term storage. Diagnostic data were available for fresh frozen tissue samples and formalin fixed paraffin embedded tissue (FFPE) from muscle biopsy specimens performed between 1994-2022 (**Table 1**). Positive patient samples and negative samples of biopsies performed for other muscular pathologies were collated, with histopathological identification performed by Anatomical Pathology, Pathology Queensland. Diagnostic histopathological features for *H.perplexum* include evidence of non-encysted nematodes measuring 350µm in length and 20µm wide in a muscle biopsy specimen. These organisms often have a tapered tail and gravid females may be present (1).

**Table 1.**
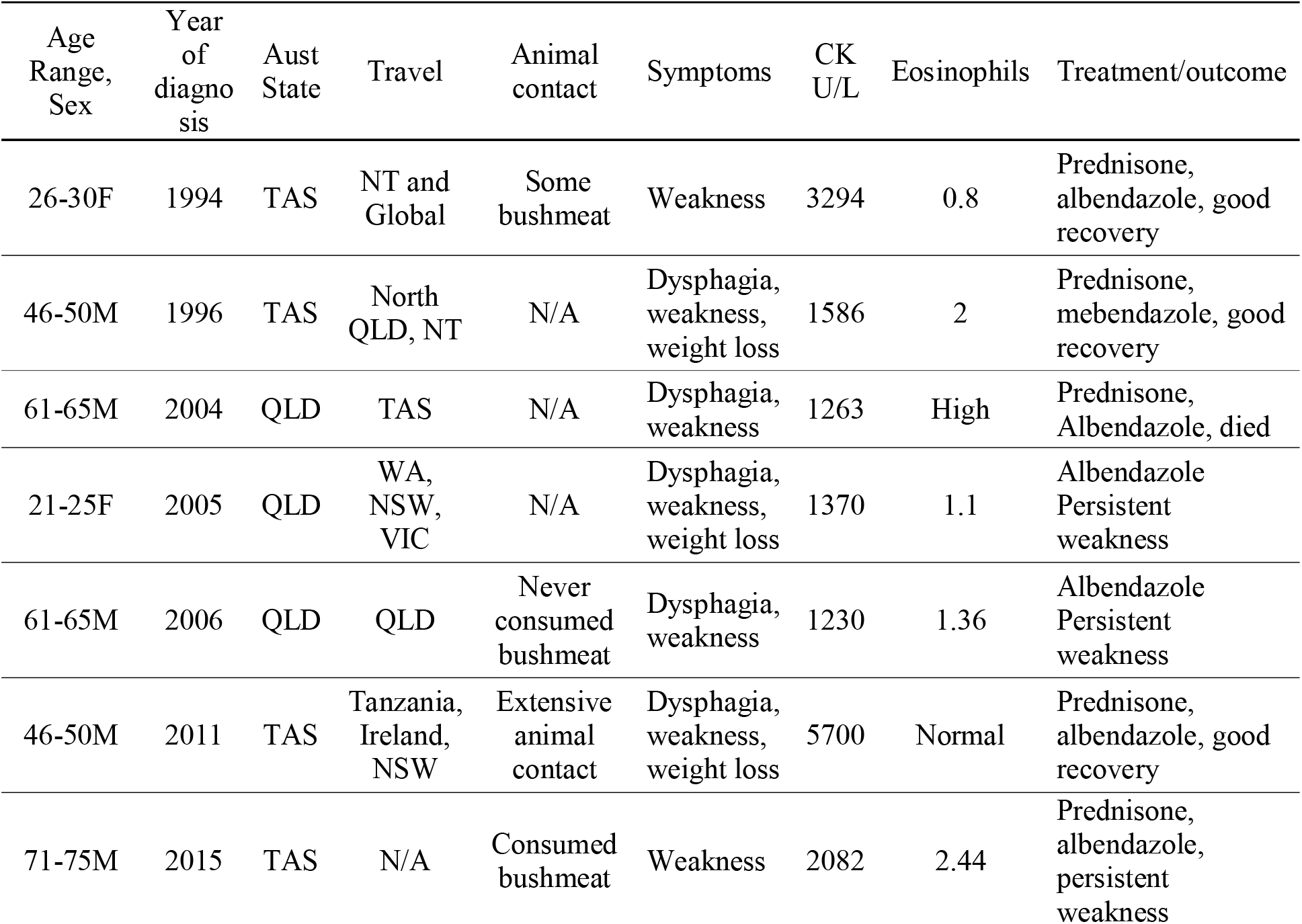
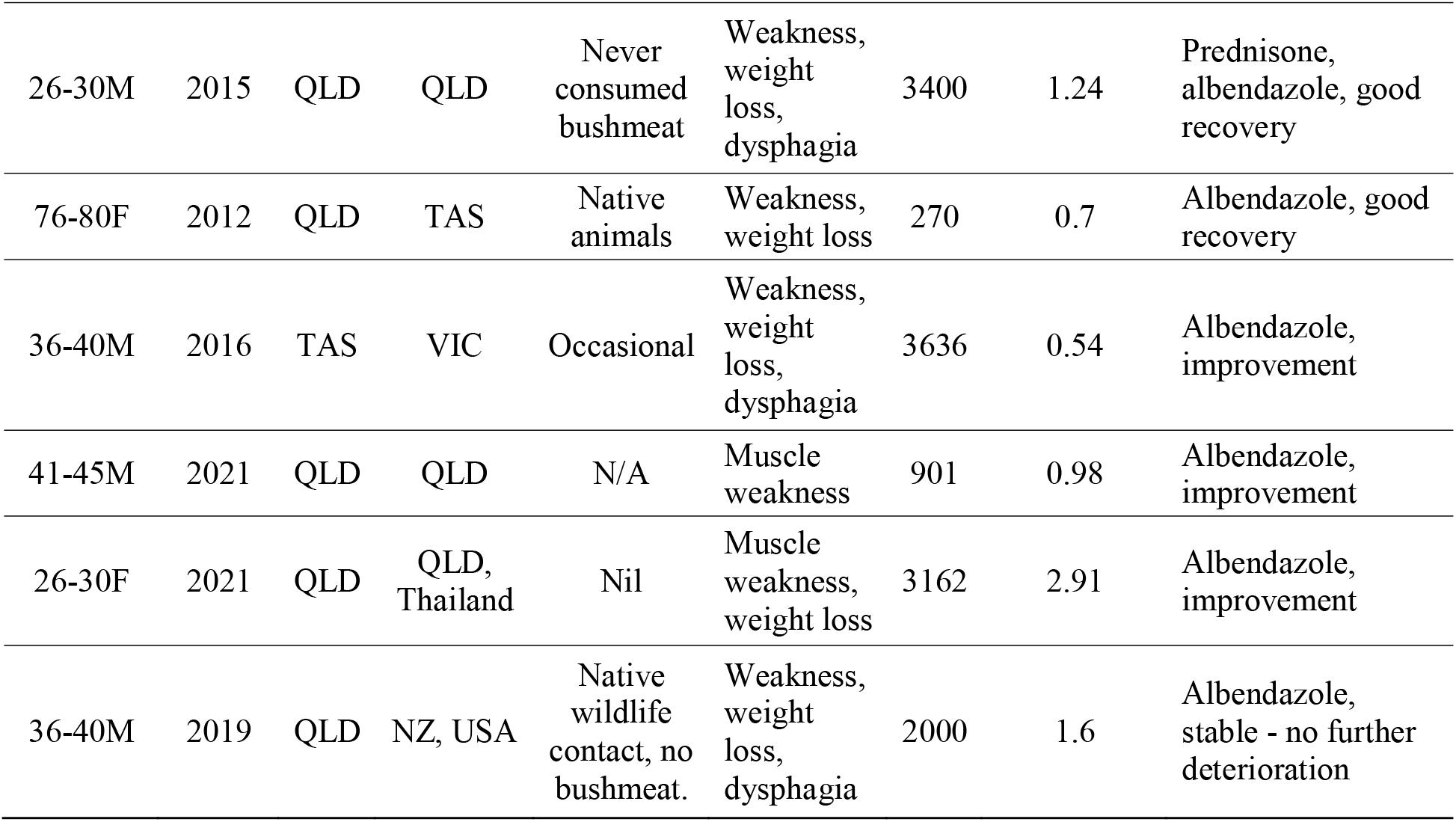
Patient and clinical characteristics of previously reported *Haycocknema perplexum* cases.

| Age Range, Sex | Year of diagnosis | Aust State | Travel | Animal contact | Symptoms | CK U/L | Eosinophils | Treatment/outcome |
| --- | --- | --- | --- | --- | --- | --- | --- | --- |
| 26-30F | 1994 | TAS | NT and Global | Some bushmeat | Weakness | 3294 | 0.8 | Prednisone, albendazole, good recovery |
| 46-50M | 1996 | TAS | North QLD, NT | N/A | Dysphagia, weakness, weight loss | 1586 | 2 | Prednisone, mebendazole, good recovery |
| 61-65M | 2004 | QLD | TAS | N/A | Dysphagia, weakness | 1263 | High | Prednisone, Albendazole, died |
| 21-25F | 2005 | QLD | WA, NSW, VIC | N/A | Dysphagia, weakness, weight loss | 1370 | 1.1 | Albendazole<br>Persistent weakness |
| 61-65M | 2006 | QLD | QLD | Never consumed bushmeat | Dysphagia, weakness | 1230 | 1.36 | Albendazole<br>Persistent weakness |
| 46-50M | 2011 | TAS | Tanzania, Ireland, NSW | Extensive animal contact | Dysphagia, weakness, weight loss | 5700 | Normal | Prednisone, albendazole, good recovery |
| 71-75M | 2015 | TAS | N/A | Consumed bushmeat | Weakness | 2082 | 2.44 | Prednisone, albendazole, persistent weakness |
| 26-30M | 2015 | QLD | QLD | Never consumed bushmeat | Weakness, weight loss, dysphagia | 3400 | 1.24 | Prednisone, albendazole, good recovery |
| 76-80F | 2012 | QLD | TAS | Native animals | Weakness, weight loss | 270 | 0.7 | Albendazole, good recovery |
| 36-40M | 2016 | TAS | VIC | Occasional | Weakness, weight loss, dysphagia | 3636 | 0.54 | Albendazole, improvement |
| 41-45M | 2021 | QLD | QLD | N/A | Muscle weakness | 901 | 0.98 | Albendazole, improvement |
| 26-30F | 2021 | QLD | QLD, Thailand | Nil | Muscle weakness, weight loss | 3162 | 2.91 | Albendazole, improvement |
| 36-40M | 2019 | QLD | NZ, USA | Native wildlife contact, no bushmeat. | Weakness, weight loss, dysphagia | 2000 | 1.6 | Albendazole, stable - no further deterioration |

### 2.2 Real-time PCR

The real-time PCR assays were developed using primers targeting both COX-1 and SSU coding regions and TaqMan probes (**Table 2**). Sequences from *H. perplexum* nematodes (GenBank accession nos. KU531719 and KU531720) were used for development of primers and probes. Samples were prepared as per protocol within the laboratory. DNA extraction was performed using the Roche MagNA Pure 96 System. The QIAgility (QIAGEN) kit and instrument was used for PCR set-up and the Rotor-Gene Q (QIAGEN) was used for PCR amplification. For both qPCR assays, SensiFast™ Probe No-ROX Mix or QuantiTect® Probe PCR Kit was used as the basis of each reaction and contained 0.5µM of forward and reverse primers, 0.2µM of FAM-labelled probe, and 5µL of nucleic acid extract in a total reaction volume of 25µL. Cycling conditions were 95°C for 15 minutes, 45 cycles of 95°C for 15 seconds and 60°C for 60 seconds and probe detection occurred via the green channel.

**Table 2.**
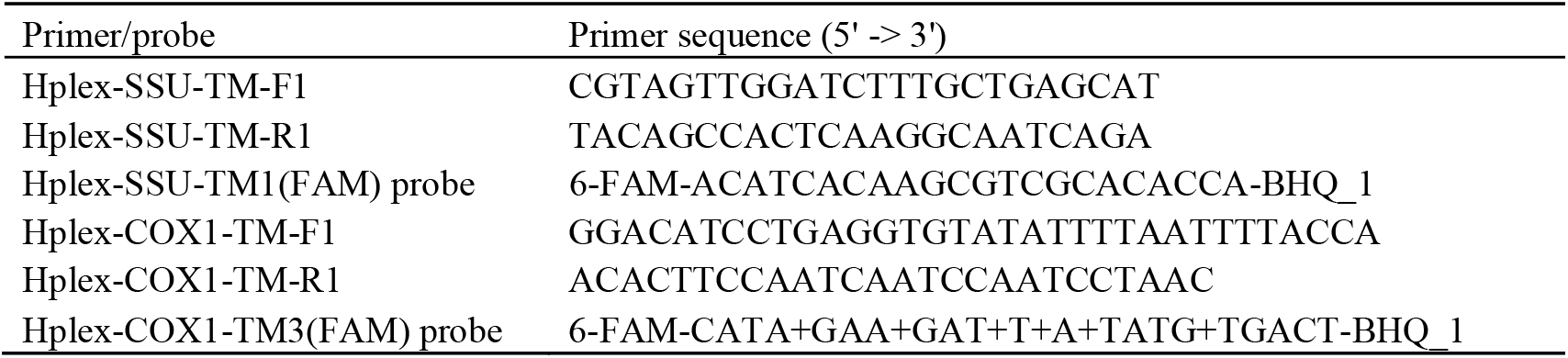
*Haycocknema perplexum* (Hplex) TaqMan qPCR primer/probes.

### 2.3 Assay performance

The performance of the assay was assessed for sensitivity and specificity. As a commercial quantified *H. perplexum* DNA control is not available for analysis of Limit of Detection (LOD), qPCR was performed using 10-fold dilutions of a sample extract DNA, reflective of a ‘real-world’ biological load, for a semi-quantitative assessment of detection at lower concentrations. Clinical specificity was determined using a sample bank of eight FFPE tissue and fresh frozen tissue specimens positive for other parasitic infections, *Enterobius vermicularis* and *Toxoplasma gondii*. These tissue biopsy samples were added to known *H. perplexum* positive tissue biopsy samples and extracted together. PCR using the *H. perplexum* single mix assays was performed on these extracts, to ensure there was no cross-reactivity with other similar microflora (**Table 2**).

### 2.4 Comparative validation of H. perplexum-extraction control duplex PCR method

A comparative validation was performed with duplexed PCRs that simultaneously detect Equine herpes virus extraction control (EHV-EC) and *H. perplexum*. Briefly, a total of 40 (12 positive and 28 negative) patient tissue samples were extracted with the addition of EHV-EC. All samples were extracted on the Roche MagNA Pure 96 system, using the automated internal control (IC) feature which aliquots 20µL of EHV-EC into each patient sample. Real-time PCR using single mix Hplex-SSU-qPCR and Hplex-COX-1-qPCR assays, EHV-single mix assay and the Hplex-SSU or Hplex-COX-1-EHV duplex mix assays were performed on these extracts (9). To ensure there was no cross-reactivity with other similar microflora, eight tissue biopsy samples positive for *Enterobius vermicularis* and/or *Toxoplasma gondii* were added to known *H. perplexum* positive tissue biopsy samples and then extracted with the addition of EHV-EC. Briefly, QuantiTect® Probe PCR Kit was used as the basis of each reaction and contained 0.5µM of forward and reverse Hplex primers, 0.2µM of FAM-labelled Hplex probe, 0.16µM forward and reverse EHV primers, 0.2µM of HEX-labelled EHV probe, and 5µL of nucleic acid extract in a total reaction volume of 25µL. Cycling conditions were repeated as above. Performance characteristics of the real-time PCR assay were calculated using histopathology as the gold-standard comparator.

## 3. RESULTS

A total of 22 samples were tested, derived from 8 patients with histopathologically-confirmed *H. perplexum* infection (**Figure 1**) This included pre-treatment diagnostic samples as well as follow-up samples post-treatment. Ten samples were tested from 5 patients without *H. perplexum* who had muscle biopsies for other muscular conditions (e.g., Inclusion body myositis, focal denervation) and 9 samples from other parasitic infections were tested for specificity. Table 3 demonstrates the results of the real-time PCR for *H. perplexum*. The assay demonstrated 100% specificity. The sensitivity of fresh frozen samples was superior to FFPE samples reporting a sensitivity of 100% for both PCR targets for diagnostic samples (pre-treatment) compared to a sensitivity of 75% for COX-1 target and 88% for SSU target for the FFPE specimen type. Overall sensitivity was 84% for COX-1 and 92% for SSU target. The quality and volume of the FFPE samples varied which may have contributed to the reduced sensitivity in this evaluation as well as multiple freeze/thaw cycles of limited nucleic acid extract. Both PCR targets were not detected on all, but one fresh specimen tested following treatment, consistent with histopathology results, demonstrating the utility of the assay for diagnosis but not monitoring.

**Table 3.** Results of evaluation of *Haycocknema perplexum* COX1-PCR and SSU-PCRs.

| Patient diagnosis | Number |  | Histopathology (N, (%) positive) | Real-time PCR |  |  | Real-time PCR |  |  |
| --- | --- | --- | --- | --- | --- | --- | --- | --- | --- |
|  |  |  |  | COX-1 (N, (%) detected) |  |  | SSU (N, (%) detected) |  |  |
|  | FFPE | FRESH | FFPE | FFPE | Fresh | total | FFPE | Fresh | Total |
| Positive Pre-treatment | 8 | 5 | 8 (100) | 6 (75) | 5 (100) | 11 (85) | 7 (88) | 5 (100) | 12 (92) |
| Positive Post-treatment | 5 | 4 | 0 (0) | 0 (0) | 0 (0) | 0 (0) | 0 (0) | 1 (25) | 1 (11) |
| Negative | 5 | 5 | Negative for *H. perplexum* | 0 (0) | 0 (0) | 0 (0) | 0 (0) | 0 (0) | 0 (0) |
| *Enterobius vermicularis* | 5 | 0 | Negative for *H. perplexum* | 0 (0) | 0 (0) | 0 (0) | 0 (0) | 0 (0) | 0 (0) |
| *Toxoplasma gondii* | 0 | 4 | Negative for *H. perplexum* | 0 (0) | 0 (0) | 0 (0) | 0 (0) | 0 (0) | 0 (0) |

**Figure 1.**
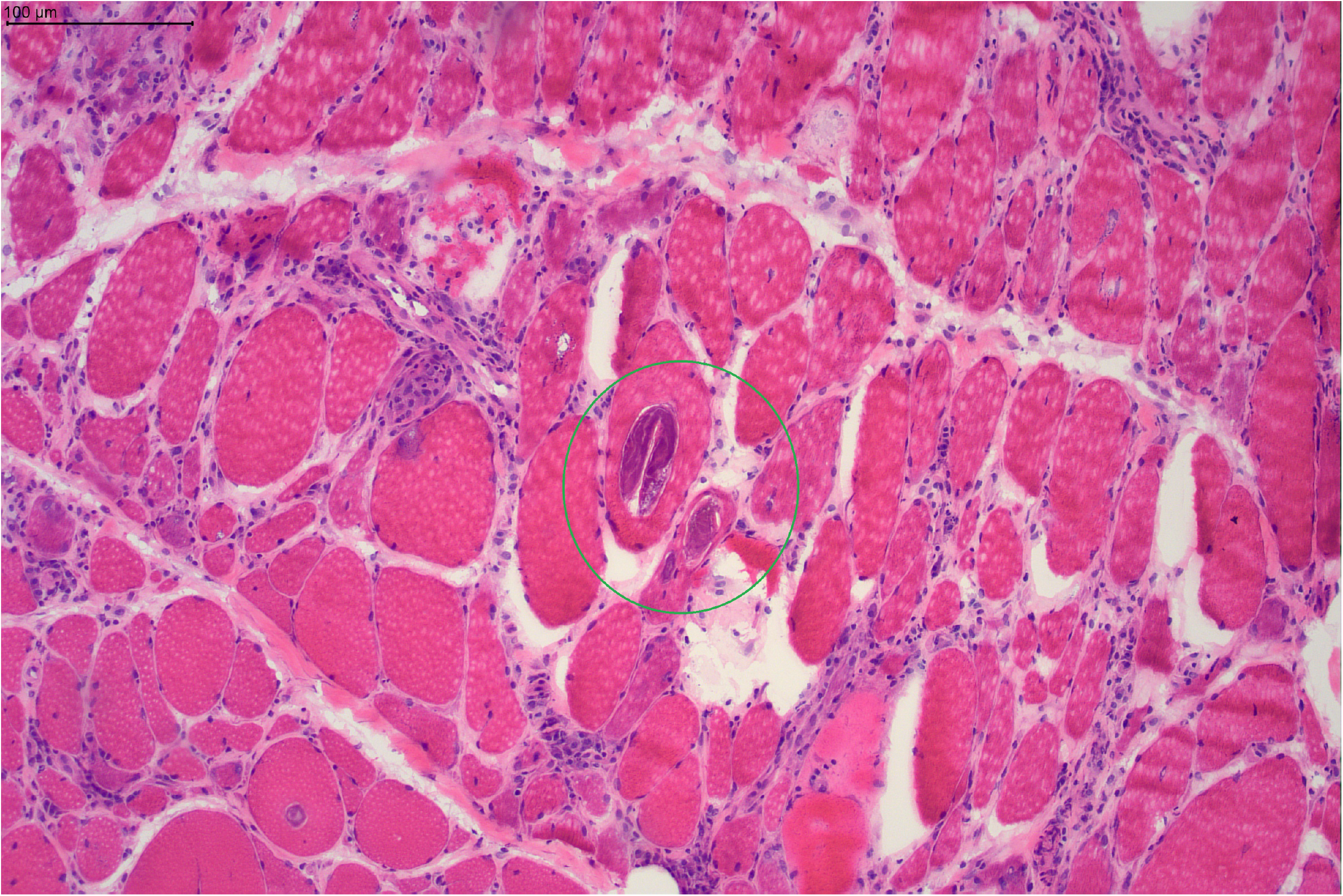
Image from haematoxylin & eosin stained frozen section through skeletal muscle biopsies from an adult male patient from Queensland, Australia. Demonstrates intra-sarcoplasmic nematodes with morphology consistent with *Haycocknema perplexum*. (Scale bar = 100µm)

A cycle threshold (C_t_) of 42 cycles was used as a cut-off for detection following a limit of detection analysis using 10-fold dilutions of both the SensiFast™ Probe No-ROX Mix and QuantiTect® Probe PCR Kits.

The mean CT detected on FFPE samples for the SSU target was 37.7, median 38.7 [range 33.9-40.8] and for the COX-1 target was 37.4, median 36.5 [range 32.7-41.]. For fresh frozen tissue, the average CT detected for the SSU target was 30.72, median 30.08 [range 28.0-34.9 and for the COX-1 target the average was 31.52, median was 31.28 [range 27.03-36.41].

The limit of detection analysis using 10-fold dilutions for the COX-1 TaqMan PCR using the SensiFast™ Probe No-ROX Mix and QuantiTect® Probe PCR Kit was 10^−4^ dilution (CT 38.15 and 40.27) and for the SSU TaqMan qPCR was 10^−3^ (CT 39.31 and 39.08). The presence of the EHV internal control was detected in all samples with consistent Ct values at between 27.68 and 30.49 for Hplex-COX-1 and between 25.99 and 29.51 for Hplex-SSU TaqMan qPCR assays, therefore when the assay was duplexed with the EHV control, the result interpretation was unchanged demonstrating no interference with detection of *H. perplexum* DNA (data not shown).

## 4. DISCUSSION

This study demonstrates the utility of this assay for diagnosis of *H. perplexum*. The sensitivity was greater on fresh frozen tissue samples (100%) in comparison with FFPE (75% COX-1, 88% SSU) highlighting the benefit of collecting fresh tissue if *H. perplexum* is suspected. However, this diagnosis is often considered retrospectively following review of histological findings. Specificity of the assay was 100% for both targets with no cross-reactivity demonstrated in this study. We have also demonstrated the ability to combine the targets for a duplex assay to enhance performance and improve workflow within the laboratory.

Of all the cases tested in this study and those previously described, all had muscle weakness with the majority also complaining of weight loss and some with dysphagia (**Table 1**). Elevated CK and peripheral eosinophilia were common laboratory findings as well as abnormal liver function tests (1). Albendazole therapy was used in most cases with varying success, 8/13 reported improvement, one reported stable symptoms and four reporting persistent weakness or further deterioration with one death in the cohort (10). Corticosteroids were used in some earlier cases and was consistently associated with worsening of symptoms and are not recommended for management.

From a diagnostic perspective, there are key histological features to aid in diagnosis and differentiate from other muscular parasites, however, one must be familiar with the organism and associated findings to identify the organism and confirm the diagnosis. The use of an adjunctive PCR could be very useful, particularly for a condition that is uncommon and still poorly understood. Our findings demonstrate good assay performance characteristics and highlight the possibility of implementing this into a laboratory workflow for testing on muscle biopsy specimens either upfront if clinically suspected or following histological examination. The use of both diagnostic modalities in conjunction is recommended. The PCR did not detect *H. perplexum* DNA in the majority of samples collected following treatment so could be used as a test of cure, however, clinical follow-up would be recommended. This assay has now been implemented into routine clinical use in our laboratory.

Efforts are underway to sequence the genome of *H. perplexum* from historical clinical specimens. Future work is still required to elucidate the taxonomy and biological characteristics of the organism and to educate and create awareness of this condition and define the optimal treatment regimen for these patients. As knowledge about this organism increases, the ability to design and evaluate less invasive diagnostic options may become possible, increasing the accessibility and likelihood of earlier diagnosis for these patients.

## Data Availability

Data are available on request

## ETHICS STATEMENT

The study was conducted in accordance with the Declaration of Helsinki and approved by the Queensland Children’s Health Services District Human Research Ethics Committee (HREC/22/QCHQ/85249), and the University of Queensland HREC (2022/HE001497). Note that two positive samples from 1994 and 1996 predated HREC approval and therefore were only included in this study for the purposes of being part of validation (quality improvement). Patient consent was waived as all samples were de-identified residual material, and this study did not involve the recruitment of patients.

## AUTHOR CONTRIBUTIONS

KL: investigation, methodology, project administration, writing – original draft, writing – review & editing. DW: methodology, resources, writing – review & editing. PH: writing – review & editing. ND: data curation, formal analysis, investigation, project administration, writing – original draft, writing – review & editing. RG: Laboratory testing and result collation. BP: Laboratory testing and result collation.

